# Long Covid active case finding: a co-produced community-based pilot within the STIMULATE-ICP study (Symptoms, Trajectory, Inequalities and Management: Understanding Long-COVID to Address and Transform Existing Integrated Care Pathways)

**DOI:** 10.1101/2022.08.24.22278954

**Authors:** Nisreen A Alwan, Donna Clutterbuck, Marija Pantelic, Jasmine Hayer, Lere Fisher, Lyth Hishmeh, Melissa Heightman, Gail Allsopp, Dan Wootton, Asad Khan, Claire Hastie, Monique Jackson, Clare Rayner, Darren Brown, Emily Parrett, Geraint Jones, Kerry Smith, Rowan Clarke, Sammie Mcfarland, Mark Gabbay, Amitava Banerjee, the STIMULATE-ICP Consortium

**Author notes:** Corresponding author: Professor Nisreen A Alwan, School of Primary Care, Population Sciences and Medical Education, Faculty of Medicine, University of Southampton, University Hospital Southampton NHS Foundation Trust, Southampton General Hospital, Tremona Road, Southampton, UK, SO16 6YD. STIMULATE-ICP Consortium Membership can be found on: https://www.stimulate-icp.org/team.

## Abstract

**Background and aim:** Long Covid is a significant public health concern with potentially negative implications for health inequalities. We know that those who are already socially disadvantaged in society are more exposed to COVID-19, experience the worst health outcomes and are more likely to suffer economically. We also know that these groups are more likely to experience stigma and discrimination and have negative healthcare experiences even before the pandemic. However, little is known about disadvantaged groups’ experiences of Long Covid and preliminary evidence suggests they may be under-represented in those who access formal care.

We will conduct a pilot study in a defined geographical area (Camden, London, UK) to test the feasibility of a community-based approach of identifying Long Covid cases that have not been formally clinically diagnosed and have not been referred to Long Covid Specialist services. We will explore the barriers to accessing recognition, care and support, as well as experiences of stigma and perceived discrimination.

**Methods:** This protocol and study materials were co-produced with a Community Advisory Board (CAB) made up primarily of people living with Long Covid. Working with voluntary organisations, promotional material are co-developed and will be distributed in the local community with engagement from key community organisations and leaders to highlight Long Covid symptoms and invite those experiencing them to participate in the study if they are not formally diagnosed and accessing care. Awareness of Long Covid and symptoms, experiences of trying to access care, as well as stigma and discrimination will be explored through qualitative interviews with participants. Upon completion of the interviews, participants will be offered referral to the local social prescribing team to receive support that is personalised to them potentially including, but not restricted to, liaising with their primary care provider and the regional Long Covid clinic run by University College London Hospitals (UCLH).

**Ethics and dissemination:** Ethical approval has been obtained from the Faculty of Medicine Ethics Committee and Research Integrity and Governance, University of Southampton. (reference number 72400). Findings will be reported in a report and submitted for peer-reviewed publication. Definitive methods of dissemination will be decided by the CAB. Summaries of the findings will also be shared on the STIMULATE-ICP website, locally in the study area and through social media. We will specifically target policy makers and those responsible for shaping and commissioning Long Covid healthcare services and social support such as NHSE England Long Covid Group.

## Background

Long Covid is a patient-coined term, initially defined as not recovering from SARSCoV2 infection after at least 4 weeks from onset (1,2). The World Health Organization defines post COVID-19 condition as the condition that occurs in those with probable or confirmed SARS CoV-2 infection, usually 3 months from onset with symptoms and that last for at least 2 months, not explained by an alternative diagnosis and impact everyday functioning (3). The Office for National Statistics (ONS) recently estimated that 2 million people are currently living with Long Covid in the UK (4). Around 12% of those infected with COVID-19 (including those initially asymptomatic) experience symptoms consistent with Long Covid for at least 12 weeks (5). Of those living with Long Covid, nearly three quarters experience limitations of their day-to-day activities, with about 1 in 5 describing their day-to-day activities as ‘limited a lot’ by Long Covid. It disproportionately affects people living more deprived areas, those with another health condition or disability, and those in frontline professions (4).

Common symptoms of Long Covid include fatigue, shortness of breath, headache, cognitive dysfunction (‘brain fog’), chest pain, muscle and joint pains, cough, disturbed sleep and psychological symptoms. These symptoms often relapse and vary at different times and can be unpredictable in nature (6,7). Evidently, Long Covid is a significant burden and public health concern. Symptoms of Long Covid can cause difficulty sustaining employment as well as increasing the need for social support (6,7). An online survey exploring the characteristics and impact of Long Covid in more than 2500 people found that 10% of people living with Long Covid had to work reduced hours, 19% reported being unable to work at all and 38% had suffered a loss of income due to their symptoms (6). The multifaceted effects of Long Covid need further exploration across different population groups.

Health disparities exist between different population groups. People from ethnic minority backgrounds or socially and economically disadvantaged people are likely to have poorer health outcomes when compared to people from a White British background or those living in more asset-rich areas (8,9). The COVID-19 pandemic has increased the visibility of these inequalities. Recent analysis shows that between January and February 2022 (when Omicron was the dominant SARS-CoV-2 variant), COVID-19 death rates were higher for ethnic minority groups compared with White British group (10). During the first and second waves, prevalence rates and mortality rates for COVID-19 were highest in ethnic minorities and those living in deprived areas (11,12). COVID-19 is also estimated to be between three and eight times more likely to result in death for people with learning difficulties compared to other adults (13,14).

Given these disparities, it is anticipated that large numbers from these communities would be needing Long Covid care and support. However, there is some evidence that individuals from less affluent and Black, Asian and minority ethnic backgrounds may be under-represented at post-covid services (15), and that primary care is not provided with additional resources to address such inequalities in diagnosis and access to care (16). To our knowledge, there is currently no evidence into how people with learning difficulties are affected by Long Covid.

Historically, disadvantaged communities have struggled with stigma and discrimination and have had bad experiences of using healthcare services (8,17). Long Covid, in itself, can be a stigmatising experience (18,19). Fear of repeating these experiences and further stigmatisation could be causing under-representation of disadvantaged groups in Long Covid primary and secondary care health services. In a survey of almost 1000 people, the authors found a prevalence of 95% and 76% of experiencing Long Covid associated stigma at least ‘sometimes’ and ‘often/always’ respectively (20). There is a need to explore the barriers to recognition and access to care in those with probable Long Covid who were unable to get a clinical diagnosis or STIMULATE-ICP (Symptoms, Trajectory, Inequalities and Management: Understanding long COVID to Address and Transform Existing Integrated Care Pathways) is a National Institute for Health Research (NIHR) supported multi-centre study. It combines a clinical epidemiological study, a multi-arm multi-site randomized controlled trial (RCT) (21) exploring the benefit of an integrated care pathway (ICP) for Long Covid, and mixed methods studies exploring inequalities of care and transferability of the ICP to other long-term conditions (LTCs) (22, 23).

This protocol forms the part of the STIMULATE-ICP study which aims to examine the inequalities in accessing Long Covid care. More specifically, this active case finding pilot, will test the feasibility of a community-based approach of identifying probable Long Covid cases that have not been clinically diagnosed and are not receiving care, whilst exploring the reasons for this and the barriers faced when attempting to obtain a diagnosis and access care and support.

## Aims and Objectives

The purpose of this pilot study is to co-design with a Community Advisory Board (CAB) and conduct an active Long Covid case-finding pilot in a specific geographical area.

The co-produced objectives with the CAB are:

a. Utilise inclusive and culturally appropriate methods to find cases not receiving recognition and care and explore referral and support pathways using a personalised approach
b. Examine the characteristics of people living with Long Covid identified through this pilot to inform clinical and social care/support pathways and learn and apply lessons around equitable service delivery
c. Develop community awareness material and healthcare professional training based on the findings
d. Provide a sampling frame for in-depth qualitative interviews with people living with Long Covid not accessing Long Covid NHS services

## Methods

This pilot study was developed using a research co-production approach with people with lived experience of Long Covid.

### Community Advisory Board (CAB)

The CAB is a Patient and Public Involvement (PPI) group. The aim was to co-produce (24) this element of the STIMULATE-ICP study assessing inequalities and stigma in Long Covid starting with the study concept and research questions (25). Co-production in research is where researchers work with patients and/or other public contributors to design and/or conduct research (24,26). Its benefits include research that is more meaningful to the population group being studied (26). The CAB was formed in partnership with our public contributor STIMULATE-ICP co-applicants (LF and JH).

LF, JH and NAA have lived experienced of Long Covid and each nominated a list of potential members of the CAB who were approached and asked if they are willing to participate in this research co-production initiative. The nominations took into account: gender, ethnicity and professional background, aiming for a balanced board composition. Twenty-nine people were approached to join the CAB through personal communication, some are members of online community support groups, and participants in JH’s Hidden Voices Long Covid Project (27). Twenty-four of those nominated and approached accepted the invitation. Some of them are health professionals as well as Long Covid patients and their professional roles were documented.

The CAB is made up of ten health care professionals (including seven who are also living with Long Covid), eight people living with or caring for children with Long Covid and five other stakeholders from voluntary organisations and public health bodies. The CAB will continue to be instrumental in the development of the active case finding until its completion.

The purpose of the CAB was discussed in the first CAB meeting and finalised during the second meeting as follows:

1. Co-design and provide advisory input on the conduct of the community-based active case finding pilot in two study sites.
2. Agree inclusion criteria for the active case-finding community-based pilot
3. Discuss appropriate ways to implement findings from the qualitative interviews and the active case-finding pilot.

To-date, three meetings have taken place. The first meeting had twelve attendees (six people living with Long Covid, five health care professionals four of whom are living with Long Covid, and one other stakeholder from the voluntary sector). Fourteen members attended the second meeting (five people living with or caring for children with Long Covid, four health care professionals, two of these living with Long Covid, and five other stakeholders from voluntary organisations and public health). The third meeting was attended by twelve CAB members (six people living with Long Covid, five health care professionals, three of whom are living with Long Covid, and one stakeholder from the voluntary sector). The first meeting centred around refining the aims of the CAB and active case finding, while in the second the methodology of the active case finding was discussed. The discussions in both meetings have fed directly into this protocol. The third meeting’s purpose was to feedback on this protocol. Discussions in the first meeting were open to capture different ideas and perspectives from the CAB members. Following meetings have become more structured as the focus of this pilot study became clearer. Notes were taken at each CAB meeting and used to aid in the development of the agenda for the following meeting. Associated study materials have been shared with CAB members. Amendments and additions to these documents have been made based on their comments.

### Active Case Finding

‘Active case finding’ can refer to the use of a range of different mechanisms in community populations to identify individuals with specific health conditions that have previously not been clinically diagnosed (28). Active case finding methodologies have previously been used to identify other illnesses including tuberculosis (28), leprosy (29) and HIV (30) within communities. The purpose of using an active case finding approach within this current study is to identify probable cases of Long Covid in the population of defined geographical areas that have not already been clinically diagnosed.

#### Location and timing of the pilot

The London Borough of Camden has some of the highest levels of inequality in England (31). Camden is a diverse borough – around 34% of people who reside there are from an ethnic minority background. There are also some areas within Camden with high levels of deprivation (32).

The research team have developed links with Camden voluntary organisations who have experience of, and are highly skilled at, identifying people in need in the community. This includes Voluntary Action Camden (VAC), who co-ordinate and deliver Community Links, for the social prescribing service in the borough and have in-depth knowledge of the locality. The research team and voluntary organisations will collaborate to identify probable cases of Long Covid in the community. In this site, this study is expected to commence August 2022 for a duration of 3 months.

#### Recruitment

An easy read leaflet has been co-developed with the CAB to raise awareness of Long Covid symptoms and functional impact in the communities being targeted for recruitment (figure 1). It describes the criteria for inclusion and what we are aiming to do. It will also form the basis of an easy read advert on community newsletters and digital outlets.

**Figure 1.**
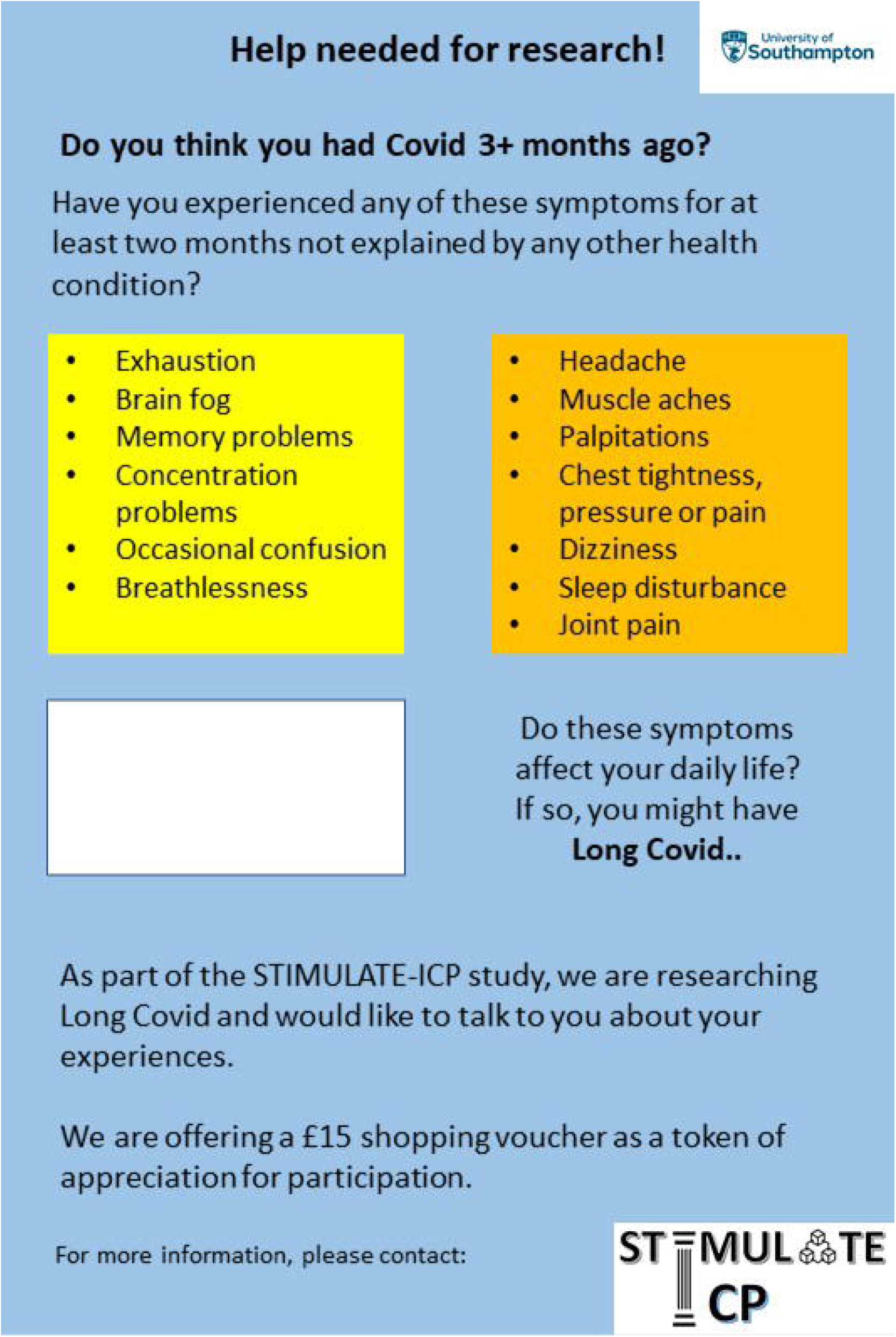
Study recruitment easy read leaflet.

**Figure 2.**
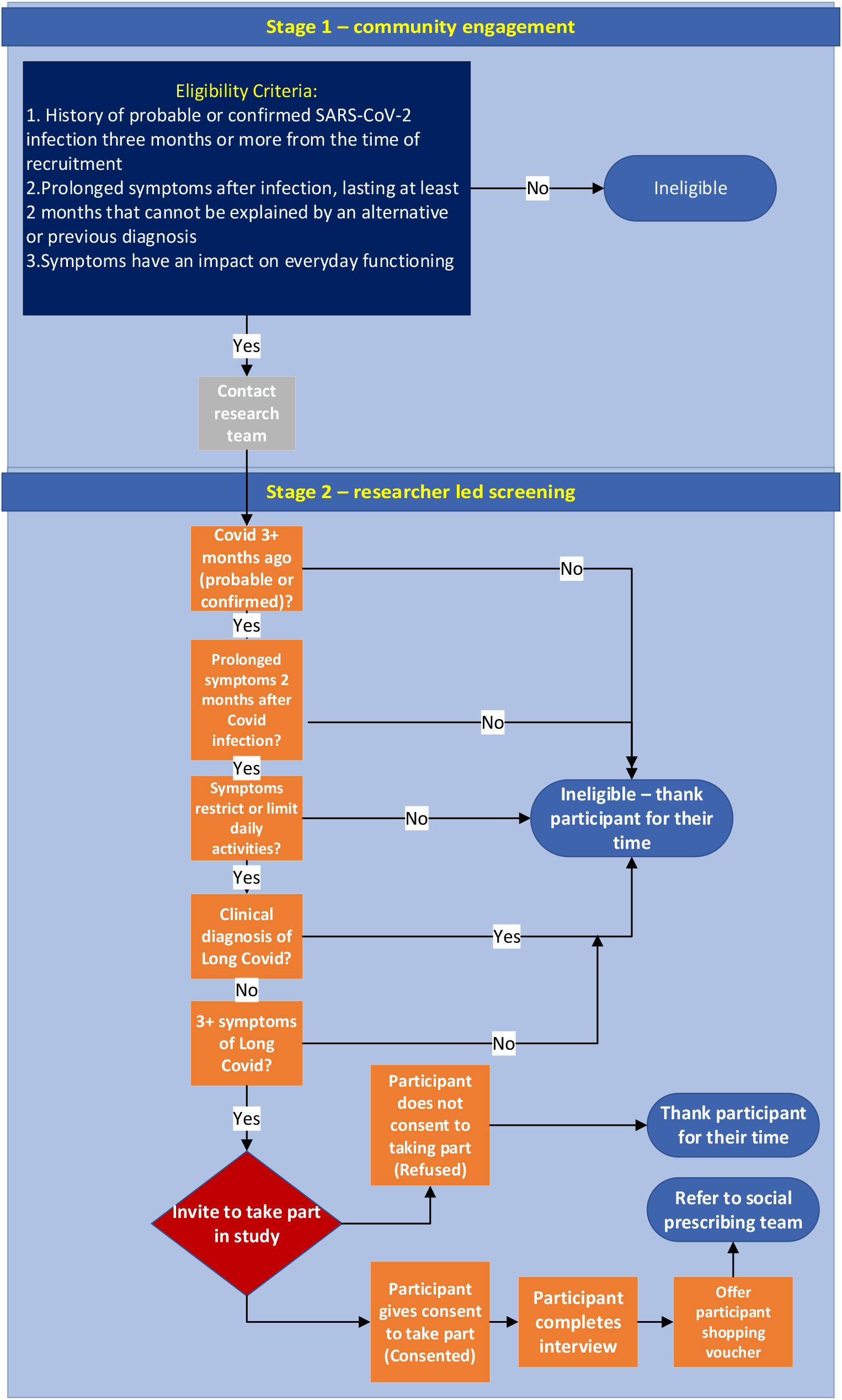
Eligibility and Screening flow chart.

The recruitment material will be further tailored to be used as educational and awareness-raising materials and to suit the target population based on the findings from this pilot study. We will seek feedback on the material as one of the study outcomes. All educational materials developed based on the findings will be reviewed by the CAB and input incorporated before finalising.

Voluntary Action Camden’s (VAC) social prescribing team will aid the research team with recruitment by distributing materials during their usual activities and in contacts with individuals referred to their service. Recruitment will also be targeted towards support groups (online and offline), social media, community venues, faith groups, colleges/universities, leaflets in pharmacies, adverts in local press or community newsletters, housing associations, food banks, resident groups in accommodation/estates.

##### Eligibility Criteria

The target population is any adult aged 18+ who lives in Camden without a clinical diagnosis of Long Covid who meets the criteria below. This includes people from ethnic minority backgrounds, individuals living in the top 20% most deprived areas in England (Index of Multiple Deprivation IMD), and individuals with (learning) disabilities. Recruiting consideration will also be given to people who are employed in frontline roles.

###### Stage 1 (reaching out)

The study’s promotional material will use the criteria from the World Health Organization Clinical Case Definition of Post COVID-19 condition to raise awareness for consideration of potential participants who may be experiencing Long Covid (1). These include:

1. History of probable or confirmed (by rapid antigen or PCR testing) SARS-CoV-2 infection three months or more from the time of recruitment
2. Prolonged symptoms after infection (a list of the 10 most common symptoms will be provided) lasting at least 2 months that cannot be explained by an alternative or previous diagnosis
3. Symptoms have an impact on everyday functioning

Potential participants who think they fulfil the above criteria will be encouraged to contact the research team by phone or email. A member of the research team will go through a set of questions for stage 2 screening below.

The 10 most common symptoms highlighted: Exhaustion/fatigue, cognitive dysfunction (brain fog, memory problems, concentration problems, occasional confusion), breathlessness, headache, muscle aches, palpitations, chest tightness/pressure/pain, dizziness, sleep disturbance and joint pain.

###### Stage 2 (screening criteria)

A screening questionnaire will be administered by a member of the research team to check eligibility. The above three criteria will be checked first. To be specific, the potential participant will be asked whether they had confirmed or probable Covid19 at least 3 months ago, whether than have experienced prolonged symptoms not explained by a pre-existing health condition for at least 2 months after infection, and whether these symptoms restrict or limits their daily activities.

If the answer is yes to all three, a question will be asked whether the potential participant has an **existing clinical diagnosis of Long Covid** (Post Covid19 Syndrome). If the answer is no, further questions about specific symptoms and their pattern will be asked. Given the evidence that Long Covid is predominantly a multi-system condition (6), participants will be included if they have **three or more** of the symptoms listed below:

Post-exertional symptom exacerbation (PESE-as defined below), exhaustion, cognitive dysfunction (brain fog, memory problems, concentration problems, occasional confusion), breathlessness, headache, muscle aches, palpitations, chest tightness/pressure, chest pain, dizziness, sleep disturbance, joint pain, leg pain, pins and needles feeling, tinnitus, sore throat, cough, nasal symptoms, hoarse voice, abdominal pain, nausea, diarrhoea, chills, altered or loss of sense of smell, altered or loss of sense of taste, skin rash, loss of appetite and sneezing.

Post-exertional symptom exacerbation (PESE): symptoms such as fatigue, difficulty thinking, pain recurring after remission or getting worse following exertion, either immediately or up to 72 hours post exertion (33).

The above screening process form the criteria for inclusion in the study rather than a tool upon which clinical decisions are made. Those who fulfil the stage 2 screening criteria will be assumed to have probable Long Covid and asked if they are interested to take part in the study. For the purposes of this pilot study, a ‘probable’ Long Covid case, is an individual who has had a confirmed or suspected SARS-CoV-2 infection and has symptoms consistent with those contained in the screening criteria above but has not received a formal clinical diagnosis.

Participants will be asked to read the Participant Information Sheet in paper or online version or a member of the research team may read it to them, or they can watch a short video describing the study using the same information included in the PIS. If they are interested to take part, they will be asked to complete the study’s consent form (in paper or online version). Arabic, Somali and Bengali are three of the most commonly spoken languages in Camden (34). If needed, Arabic, Somali and Bengali versions of the PIS short video can be provided. Study documents can be translated into these community languages if this is required following an assessment of recruitment progress 6 weeks into the study. Some participants, especially those with additional needs or those who speak community languages, may prefer to have the documents read and/or explained to them by a friend, carer or voluntary organisation worker. Whether they opt in the study or not, they will be directed to NHS resources and/or NHS 111 website for further information about the kind of help they can seek regarding their symptoms.

#### What happens after cases have been identified and they have consented to take part?

##### Qualitative research data collection

Participants will be invited to take part in a qualitative semi-structured interview conducted by a member of the research team. The qualitative interview will explore participants’ awareness of Long Covid and symptoms, their experiences of trying to access care and barriers, as well as stigma and perceived discrimination by asking broad, open questions. Example questions within the interviews: ‘Please can you tell me a bit about your experience of Long Covid?’; ‘How comfortable have you felt seeking Long Covid care?’ ‘How have you tried to seek Long Covid treatment or care’; ‘What do you think could make it easier for people to access Long Covid care?’; ‘what do you think there is anything (outside of health care) that could make it easier for people living with Long Covid?’ Prompts may sometimes be used to encourage participants to give more full descriptions of their experiences. Prompts will include reference to: symptoms and whether these have been relapsing and/or episodic in nature (6,35); different forms of care/support sought; barriers and perceived discrimination when accessing care; and different forms of support.

Stigma questions within the topic guide are based on a validated Long Covid stigma scale (20) based on health stigma theory, emerging qualitative evidence of Long Covid stigma and stigma questions used for other conditions such as HIV and myalgic encephalomyelitis.. The semi-structured nature of the interviews means that the research team use a topic guide to inform the areas to discuss during the interview. There is no expectation to rigidly follow any order of questions instead it is a guide of areas of interest with open questions about experience and opinions with some prompts for the interview of specific areas of particular relevance to the research questions. The researcher will be able to adapt the interview to fit each participant, depending on the participant’s answers. Participants will also be asked for feedback on the pilot’s promotional material that they have accessed and input on what is important for them to see in future educational material on Long Covid.

Interviews will take place via video call, telephone or face-to-face. Participants who are unable to complete the interview via these formats will be able sent a written version of the topic guide and invited to write their own responses and add additional areas relevant and important to them. Interviews will take around 45-90 minutes to complete, depending on the participants’ answers. It is expected that 20-30 interviews will need to be completed before data saturation occurs. Some participants may prefer a friend, carer or voluntary organisation representative to be present to make the process easier for them.

Video and telephone interviews conducted through MS Teams will be digitally recorded, transcribed by the software., then checked through by the interviewer. Face to face interviews will be recorded and then transcribed by a professional transcription service. Once an interview has been transcribed, the digital recording will be deleted. Transcripts will be stored securely on University servers.

Thematic analysis will then be used to analyse the transcripts. Thematic analysis will be based on the approach set out by Braun and Clarke (36). This approach involves becoming familiar with the interview transcripts, creating codes based on participants’ responses and in relation to the research questions, creating themes that emerge from the transcripts and checking and revising these themes (if necessary) before writing the results (36). This analysis will be aided by NVivo 12 qualitative analysis software.

### Research Participation Reimbursement

Participants who complete the qualitative interview will be offered a £15 paper or online shopping voucher as a gesture of recognition for their time. This decision was based on CAB members unanimously agreeing that a financial incentive would be necessary to show gratitude to participants who take part in interviews. The research team do not believe that any ethical issues will arise from offering this voucher incentive; the monetary value of the voucher is too low to be considered coercive (37). The intention behind the payment offer is to show that the participants’ time and effort are valued.

### Community care pathway

During our CAB discussion, accessing Long Covid clinics/services was challenged as the only end goal for most people living with Long Covid, in contrast to a person-centred approach based on identified needs instead. A personalised approach could be piloted in service provision, including clinical and non-clinical pathways and social support. Improving health outcomes is a priority, but individuals with Long Covid may need help solving other problems, such as debt advice or housing support. The feasibility of a multi-pronged community-based service approach will be explored by our local partners as described below.

VAC is an ‘umbrella’ organisation that links with and supports other voluntary organisations in the Camden area (38). VAC works with the North Central London (NCL) Care Commissioning Group (CCG) and Age UK Camden to provide The Care Navigation and Social Prescribing Service in the local area (39). The aim of social prescribing is to improve mental and physical health outcomes by signposting and referring individuals to services that can make differences to health. Social prescribing recognises that physical and mental health outcomes can be influenced by ‘social, economic and environmental factors’ and aims to provide support for individuals that take these different elements into consideration (40). Furthermore, some evidence suggests that these improvements in health outcomes can be made through social prescribing (41).

Residents in Camden can self-refer for the local social prescribing service, but referrals also come from health services (39). VAC have recently began receiving referrals from the University College Hospital (UCLH) post-covid clinic as part of a pilot pathway to see if social prescribing can have a positive impact on patients living with Long Covid.

Social prescribing will be incorporated into the pilot at the Camden site. Once the qualitative interview has been completed, consent will be sought from the participant to refer them to VAC social prescribing team (if they have not accessed the study through signposting from VAC already). Social prescribers will speak with the participant if they consent and develop a person-centred approach to provide support that is meaningful to them. This might be a healthcare-focused approach but social prescribers can also make referrals to different organisations such as debt help, help with budgeting and peer support, amongst others (39). As some participants will be recruited through signposting from VAC, it is likely that some participants may have these conversations with social prescribers prior to contact with the research team. This element will be part of routine local service delivery rather than research.

Social prescribers will be able to send a referral through to University College Hospital (UCLH) Post-Covid service if necessary. The participant will be advised to contact their primary care provider to discuss their symptoms and consider a clinical assessment and/or an e-referral to the post-covid service through the electronic system if deemed appropriate by their GP. Following consent of participants, the study team can also provide a letter for the participant to forward to their GP once the qualitative interview is completed if requested by the participant. If the social prescribing team is involved, they may also contact the GP as part of service provision if advocacy is required. The participant’s wait-time for the UCLH clinic will begin from when the social prescribers send through the referral if deemed appropriate within local and national guidelines. Social prescribers will also be able to refer patients to the Camden Borough level Multi-disciplinary Team (MDT) to be put on their meeting schedule if deemed appropriate.

All identified cases will also be provided with a list of local support organisations. If the need for more urgent referral is needed, the participant will be strongly encouraged to promptly contact their GP or NHS 111 as appropriate.

#### Outcomes

The measures of success and key outcomes of this pilot study will be:

- Co-produced appropriate Long Covid educational/awareness materials that have received input during the pilot
- 20-30 people with probable Long Covid participants recruited
- Described characteristics of people identified within this pilot who are not accessing care
- Described the barriers to accessing care and the stigma experienced by people living with Long Covid
- Piloted a personalised referral/support pathway as part of service delivery
- Developed components of health professional training based on the findings of this active case finding to enhance understanding of the experiences of people living with Long Covid
- Advising local organisations on Long Covid plans that are locally specific

### Ethics and risk management

This pilot forms part of the NIHR-funded STIMULATE-ICP study (42). This includes three work-packages. This protocol refers to work under work-package 3a. Work-package 2 of STIMULATE-ICP is a pragmatic cluster randomised trial (IRAS project ID 1004698) (21). WP1 and WP3 are the observational elements (IRAS project ID 303958) (22). Participants for this pilot will be recruited from the community not the NHS and therefore NHS IRAS/REC approval was not sought for this element. The active case finding study protocol and related study documentation have received ethical approval from the University of Southampton Faculty of Medicine Ethics Committee and the Research Integrity and Governance team (reference number 72400). Informed consent will be obtained from all participants prior to their participation in this pilot study.

### Publication and dissemination

Findings will be reported in a report and submitted for peer-reviewed publication. Definitive methods of dissemination will be decided by the CAB. Summaries of the findings will also be shared on the STIMULATE-ICP website, locally in the study area and through social media. We will specifically target policy makers and those responsible for shaping and commissioning Long Covid healthcare services and social support.

## Discussion

Long Covid is a public health concern with increasing evidence of significant burdens on the lives of those living with it. This burden is complex and goes beyond the direct health consequences to social and economic implications, which in turn affect health. Health inequalities in England exist, with different groups experiencing worse outcomes from COVID-19. Little is known about the effects of Long Covid on these groups. More research is needed to investigate the causes of inequalities in Long Covid care and outcomes, as well as potential ways to reduce such health inequalities and provide evidence to inform any changes to the system towards being more receptive to the needs of underserved ethnic and more socially deprived population groups.

This active case finding sub-study of STIMULATE-ICP will identify probable Long Covid cases who are not accessing clinical care for Long Covid whilst exploring the barriers, stigma and perceived discrimination individuals with Long Covid symptoms may face, whether this is through their attempts to obtain support or through their daily life activities and societal interactions.

The main strength of this study is the co-produced nature of its design. We anticipate that the CAB’s involvement from the beginning of the study would result in findings that are relevant to people living with Long Covid. The framing of research questions and subsequently the design and methods with people with lived experience is a positive step towards avoiding past mistakes in relation to post viral illness research that does not address the main issues patients face in relation to healthcare, social care and other societal implications of their disability. If research questions are biased against the everyday experiences patients face, the answer will not be useful to improve practice (25).

It is hoped that findings from this study will help enhance understanding and inform ways through which disadvantaged communities are able to access appropriate and meaningful Long Covid care and support, including providing evidence towards a more meaningful and effective patient-clinician interaction around Long Covid symptoms and the disability they cause. The findings can build on already existing literature addressing how to leverage the patient-professional communication to optimise symptom management and reduce the extra disadvantage caused by them (43).

## Data Availability

This is a study protocol with no associated data at this stage.

## Acknowledgements

Our sincere thanks to all of those who contributed during the Community Advisory Board meetings including Rachael Gosling, Andrew Lee, Donna Turnbull and Aleyah Babb-Benjamin. This protocol is part of the STIMULATE-ICP study including Paula Lorgelly, Elizabeth Murray, Hakim-Moulay Dehbi, Hugh Montgomery, Sarah Clegg, Henry Goodfellow, Mel Ramasawmy, Yi Mu, Toby Hillman, Emma Wall, Michael Zandi, Caroline Watkins, Denise Forshaw, Gordon Prescott, Gregory Lip, Dan Cuthbertson, Nefyn Williams, Michael Crooks, Angela Green, Christina van der Feltz-Cornelis, Jenny Sweetman, Han-I Wang, Natalie Smith, Kamlesh Khunti, David Strain, Chris Robson, Mike Brady, Rajarshi Banerjee, Cat Kelly and Emily Attree. An up-to-date version of Consortium members can be found on https://www.stimulate-icp.org/team. STIMULATE-ICP can be contacted at

## Author Contributions

Conceptualisation: NAA, MP, MG, GA, AB, JH, LF

Design and methodology: NAA, DC, MP, JH, LH, AK, CH, CR, DB, EP, GJ, KS, RC, SM, DW, MH

Writing: NAA, DC

Review and/or editing: all

## Open Access

For the purpose of open access, the author has applied a Creative Commons Attribution (CC BY 4.0) licence to any Author Accepted Manuscript version arising.

